# Spike vs nucleocapsid SARS-CoV-2 antigen detection: application in nasopharyngeal swab specimens

**DOI:** 10.1101/2021.03.08.21253148

**Authors:** Moria Barlev-Gross, Shay Weiss, Amir Ben-Shmuel, Assa Sittner, Keren Eden, Noam Mazuz, Itai Glinert, Elad Bar-David, Reut Puni, Sharon Amit, Or Kriger, Ofir Schuster, Ron Alcalay, Efi Makdasi, Eyal Epstein, Tal Noy-Porat, Ronit Rosenfeld, Hagit Achdout, Ohad Mazor, Tomer Israely, Haim Levy, Adva Mechaly

## Abstract

Public health experts emphasize the need for quick, point-of-care SARS-CoV-2 detection as an effective strategy for controlling virus spread. To this end, many “antigen” detection devices were developed and commercialized. These devices are mostly based on detecting SARS-CoV-2’s nucleocapsid protein. Recently, alerts issued by both the FDA and the CDC raised concerns regarding the devices’ tendency to exhibit false positive results. In this work we developed a novel alternative spike-based antigen assay, comprised of four high-affinity, specific monoclonal antibodies, directed against different epitopes on the spike’s S1 subunit. The assay’s performance was evaluated for COVID-19 detection from nasopharyngeal swabs, compared to an in-house nucleocapsid-based assay, composed of antibodies directed against the nucleocapsid. Detection of COVID-19 was carried out in a cohort of 284 qRT-PCR positive and negative nasopharyngeal swab samples. The time resolved fluorescence (TRF) ELISA spike-assay displayed very high specificity (99%) accompanied with a somewhat lower sensitivity (66% for Ct<25), compared to the nucleocapsid ELISA assay which was more sensitive (85% for Ct<25) while less specific (87% specificity). Despite being out-performed by qRT-PCR, we suggest that there is room for such tests in the clinical setting, as cheap and rapid pre-screening tools. Our results further suggest that when applying antigen detection, one must consider its intended application (sensitivity vs specificity), taking into consideration that the nucleocapsid might not be the optimal target. In this regard, we propose that a combination of both antigens might contribute to the validity of the results.

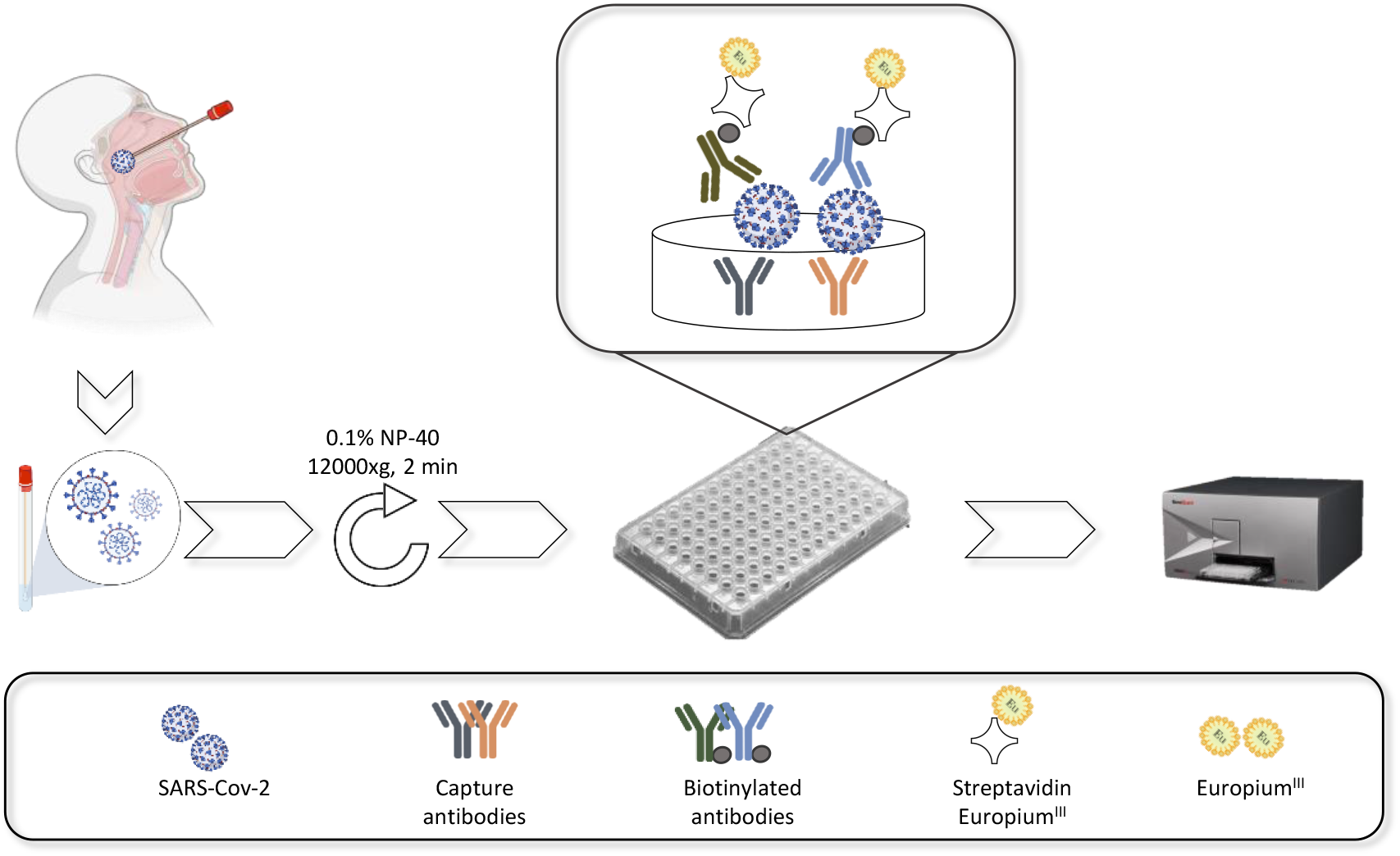

**Graphic abstract:** Schematic representation of sample collection and analysis. The figure was created using BioRender.com

## Introduction

In December 2019, a novel zoonotic coronavirus (SARS-CoV-2), identified initially in Wuhan China, led to the pandemic known as COVID-19 (1). While some affected individuals are asymptomatic, others exhibit mild symptoms such as fever and cough with some deteriorating to pneumonia, severe acute respiratory syndrome and death (2, 3). According to recent updates (February 2021), the estimated death toll for this world-wide pandemic, is about 2M people with over 100M verified cases (https://www.worldometers.info/coronavirus/).

With FDA approved vaccines emerging only recently (Pfizer and Moderna RNA-based and Oxford-AstraZeneca viral-based vaccines, December 2020), the primary tools for limiting the spread of the disease and driving down the basic reproduction number (R0) are diagnosis, surveillance and quarantine (4). The gold standard for COVID-19 diagnosis is nucleotide-based testing (qRT-PCR) of viral RNA in nasopharyngeal swabs, collected from the upper respiratory tracts of suspected individuals. This test is sensitive and specific, but is both expensive and time consuming. The gap between the ability to perform qRT-PCR in a timely manner and the need to apply rapid containment of infected individuals is a major limitation, preventing fully effective curtailing of infection chains, a key factor in disease control. There is therefore a critical demand for alternative, rapid diagnostic tests which are cheap and easy to perform and can serve for high throughput routine screening and/or triage of infected individuals at the doctor’s office (point-of-care) or in the community. Accordingly, many antigen diagnostic kits were developed, some of which were emergency-approved by the FDA and are now commercially available for use for SARS-CoV-2 detection (5).

Most of the FDA approved commercial antigen kits (12 out of 13) target the nucleocapsid (NC). This protein, enclosed within the viral lipid membrane, is one of four structural proteins encoded by the SARS-CoV-2 positive-sense RNA viral genome. On November 3^rd^ 2020, the CDC issued a lab alert, expressing concerns regarding a potential for false positive results when implementing antigen tests for rapid detection of COVID-19, especially in the community (6). A similar warning was issued by the FDA (7). While this might be an inherent limitation of antibody-based tests, it could possibly be mitigated by targeting a different antigen. One such potential target is the spike protein (S). This protein is part of SARS-CoV-2 outer membrane (8) and is responsible for the “crown” like protrusions that are a characteristic of corona viruses. There are several publications demonstrating SARS-CoV-2 detection via its spike protein (9-17). However, most only demonstrate detection of the spike protein itself or implement virus-like particles (9-15, 17). Only one work (16), applying a highly sensitive field-effect transistor-based biosensor, showed direct SARS-CoV-2 detection from swab specimens of three hospitalized patients. None of the above-mentioned assays, address the issue of sensitivity and specificity in a large cohort of nasopharyngeal swabs.

In this work we wanted to assess the potential use of the spike protein as a target for SARS-CoV-2 detection from nasopharyngeal swabs. To this end we implemented our recently developed monoclonal anti-spike antibodies (18, 19) in an in-house antibody-based test for SARS-CoV-2 detection via its spike protein. For comparison, we developed an additional in-house test, targeting the nucleocapsid. The sensitivity and specificity of both tests were evaluated for detection of COVID-19 in a large cohort of nasopharyngeal swabs of both symptomatic and asymptomatic qRT-PCR positive and negative individuals.

## Materials and Methods

### Antigens and antibodies

Nucleocapsid – SARS-CoV-2 nucleocapsid phosphoprotein’s sequence (GenBank accession: YP_009724397.2) was codon-optimized for *E. coli* expression, synthesized and cloned (*EcoRI, NotI*) in pET-28a(+) expression vector, by twist bioscience (twist bioscience, USA). The Tobacco Etch Virus (TEV) Protease consensus sequence was added downstream to the 6xHis-tag coding sequence in the NC C-terminus. The resulting plasmid DNA was maintained in a T7 Expressing lysY Competent *E. coli* strain (New England Biolabs, France) and the transformed bacteria were cultured in terrific broth (100 μg/ml ampicillin, 250 rpm, 37°C). For protein expression an overnight culture was adjusted to 0.1 optical density (at O.D 600nm) in the same medium, induced (0.1 mM IPTG at 1 O.D 600) and incubated overnight at 15°C. Cell pellet was harvested and lysed using Bugbuster® master mix lysis buffer (merckmillipore, USA) according to the manufacturer’s protocol. Urea 8M was added to the pellet and the NC protein was purified using HIS-Select®Nickel affinity gel (Sigma-Aldrich, USA) according to the manufacturer’s instructions.

Spike - A stabilized soluble version of the SARS-CoV-2 spike protein was designed by inclusion of proline substitutions at positions 986 and 987, and disruptive replacement of the furin cleavage site RRAR (residues at position 682-685) with GSAS, as reported (20). The SARS-CoV-2 spike glycoprotein (s2p) and S1 subunit were generated as previously described (18).

SARS-CoV-2 virus was cultivated as described in (21) on VERO E6 cells. SARS-CoV-2 (GISAID accession EPI_ISL_406862) was provided by Bundeswehr Institute of Microbiology, Munich, Germany. Handling and working with SARS-CoV-2 virus were conducted in a BSL3 facility in accordance with the biosafety guidelines of the Israel Institute for Biological Research (IIBR).

Antibodies: Polyclonal anti NC protein antibodies were prepared by immunizing rabbits with 100μg of purified protein in incomplete freund’s adjuvant by subcutaneous injection. The vaccination regime included a prime and two boost doses, given in 4-week intervals. Serum was collected 10 days post the second boost. Monoclonal anti SARS-CoV-2 RBD, NTD and NC antibodies were isolated from a phage-display library, constructed from COVID-19 patients showing substantial illness and panned against plate adhered spike and nucleocapsid as previously described (18, 19, 22). Antibodies were expressed and purified as described (18).

### Antibody labeling

Biotinylation of IgG purified antibody fractions was carried out using sulfo-NHS-SS-biotin [sulfosuccinimidyl-2-(biotinamido) ethyl-1,3-dithiopropionate; Pierce 21331] according to the manufacturer’s instructions. A calculated average number of 4 biotin molecules per antibody was determined by the HABA ([2-(4-hydroxyazobenzene] benzoic acid) method (Pierce 28050).

### Biolayer interferometry for affinity measurements and epitope binning

Binding studies were carried out using the Octet system (ForteBio, USA, Version 8.1, 2015) that measures biolayer interferometry (BLI). All steps were performed at 30°C with shaking at 1500 rpm in a black 96-well plate containing 200μl solution per well. For affinity measurements, streptavidin-coated biosensors were loaded with biotinylated antibodies (5 µg/ml) to reach 1 nm wavelength shift followed by a wash. The sensors were then reacted for 300 seconds with increasing concentrations of RBD/NTD (association phase) and then transferred to buffer-containing wells for another 600 seconds (dissociation phase). Binding and dissociation were measured as changes over time in light interference after subtraction of parallel measurements from unloaded biosensors. Sensorgrams were fitted with a 1:1 binding model using the Octet data analysis software 8.1 (Fortebio, USA, 2015), and the presented values are an average of several repeated measurements (At least two repeats). For binning experiments of antibody pairs, antibody-loaded sensors were incubated with a fixed S1 concentration (200 nM), washed and incubated with the non-labeled antibody counterpart. For concomitant binding, antibodies were consecutively loaded on the pre-exiting antibody-antigen complex, with no regeneration step.

### Assay development

The developed assay was a 3-step sandwich ELISA, based on Time Resolved Fluorescence (TRF). A 50µl solution of the capture antibodies (in 50mM Na2CO3, pH 9.6) was added to the microplate wells and incubated at 4°C O.N. The plates were then washed (3xPBS+0.05% Tween20) and blocked (100µl/well, with PBS containing 2%BSA and 0.05% Tween20). After one hour of incubation at 37°C the plates were washed (3xPBS + 0.05% Tween 20) and stored at −20°C or used directly. Samples (proteins or virus) were diluted in PBS or nasopharyngeal swabs buffer, and loaded on the plates. Nasopharyngeal swabs samples were added to the plate following addition of NP-40 (to a final concentration of 0.1%) and centrifugation (2 min, 15000 rpm, 4°C). After 30 minutes incubation the plates were washed and biotinylated reporter antibodies were loaded for additional 30 min. After an additional wash step, streptavidin-europium diluted 1:1000 was loaded for 20 min. After a final wash enhancement solution (Delfia, Perkin elmar) was added and the resulting signal was measured using a microplate reader (excitation 340nm, emission 612nm).

### Clinical samples

Nasopharyngeal swab specimens were collected as part of routine nursing home sampling in Israel (tenants and staff) and from the emergency department of SHEBA Medical Centre (SMC, Ramat Gan, Israel). qRT-PCR testing of samples collected from SMC was carried out in SMC while the nursing home samples were tested in-house. Samples from SMC were accompanied with clinical details regarding symptomology, as well as time elapsed since symptom onset. A total of the 46 positive and 23 negative qPCR samples were obtained from the emergency department and a total of 100 positive and 115 negative qPCR samples were acquired from the nursing homes. Samples were analyzed by the in-house developed antigen tests (Not all samples were analyzed by both tests due to insufficient sample volume). The study was approved by the SMC institutional review board committee (approval number – 7769-20-SMC).

### Signal analysis

TRF signals were calculated as signal-to-noise (S/N) ratios between the fluorescence values of antigen containing samples (‘S’ for signal) - or PBS/swab buffer containing samples (‘N’ for noise). To determine the limit of detection (LOD) for each test, the average background fluorescence was calculated as the mean of at least six “noise” replicates (PBS/swab buffer). The LOD was defined as 3 standard deviations (SD) above the average background with a coefficient of variation (CV) < 20%. These values were used to calculate the signal-to-noise ratio that was used as a cutoff value (S/N ratios ≥ 1.7) and was considered to be the LOD threshold in accordance with ICH guidelines for validation of analytical procedures (23). This calculation enabled the normalization of multiple experiments. Sensitivity was defined as the proportion of the PCR positive samples that tasted positive in the ELISA tests while specificity was defined as the proportion of PCR negative samples that tasted negative in the ELISA tests.

## Results

### Development of a TRF ELISA test for detection of SARS-CoV-2 spike protein

We recently isolated a panel of high-affinity, anti-spike human monoclonal antibodies from phage-display libraries generated from sera of COVID-19 patients (18, 19). The performance of these antibodies was evaluated for SARS-CoV-2 detection in a TRF ELISA-based format. The optimized spike assay (S-assay) includes a combination of four monoclonal antibodies that bind separate epitopes and are thus able to bind simultaneously to the spike protein with no apparent structural hindrance as determined using BioLayer interferometry - BLI (Fig 1a). Three of the antibodies (BL6, MD29 and MD65) recognize the RBD region of the spike protein, while the additional antibody (BL11) recognizes the NTD region, (represented schematically in Fig 1b). The affinities of the incorporated antibodies (determined using BioLayer interferometry) range from 0.4 to 2.5nM (Fig 1c and (18)). In the final format of the S-assay the anti-NTD antibody (BL11) and one of the anti-RBD antibodies (BL6) are incorporated as capture antibodies and the two remaining anti-RBD antibodies (MD29 and MD65) as reporter antibodies.

**Fig. 1.**
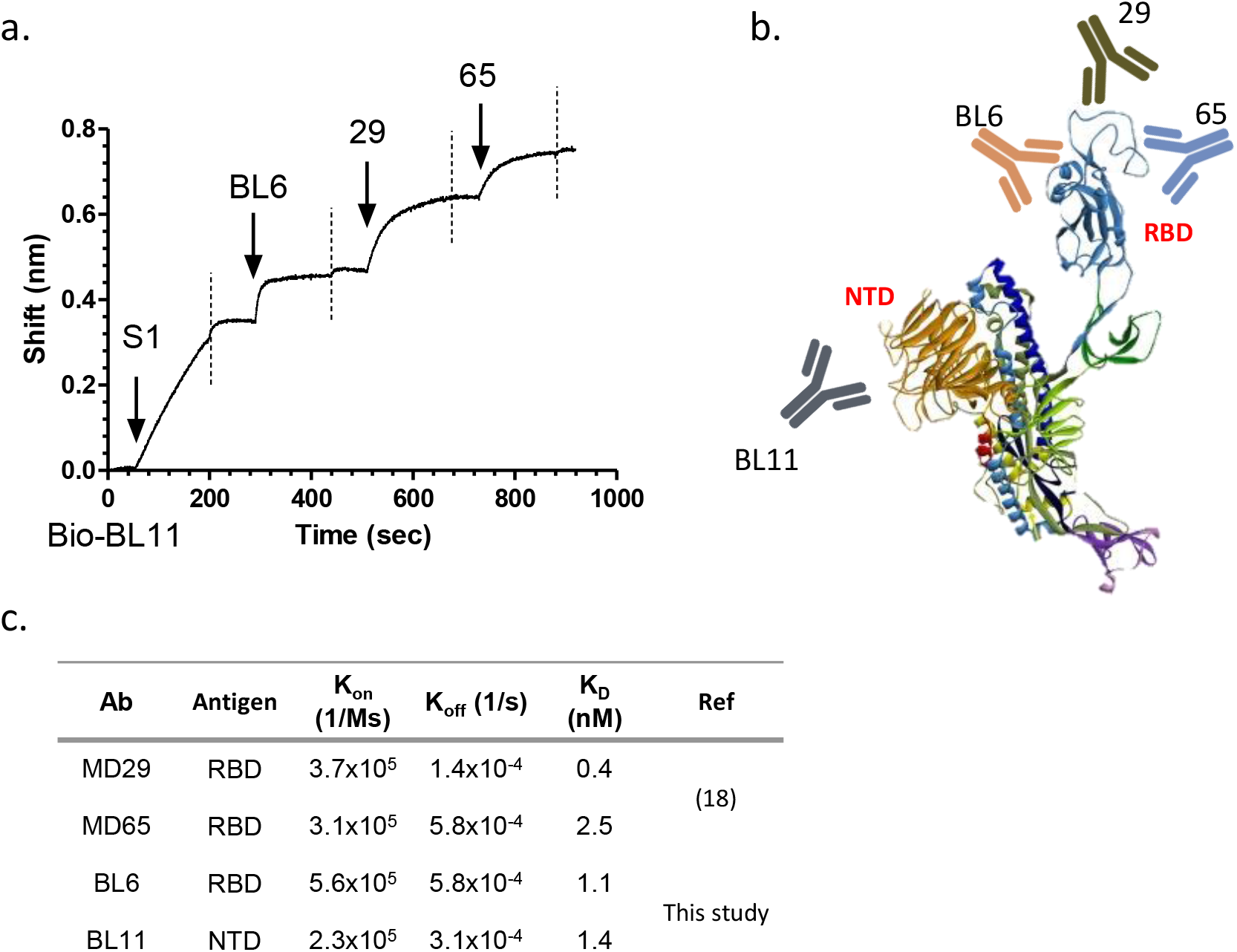
Characterization of the antibodies incorporated in the SARS-CoV-2 TRF-based S-assay. a. Concomitant binding of anti-SARS-CoV-2 antibodies. The ability of the assay’s antibodies to bind simultaneously to SARS-CoV-2 was tested using the Octet Red biolayer interferometry (BLI) system. Biotinylated BL11 was immobilized to a streptavidin sensor and interacted with the spike’s S1 subunit. The complex was then immersed in a well containing the indicated antibody (pointed by an arrow), washed again (dashed vertical line) and immersed with the next antibody. b. Schematic representation of the interaction of the assay’s antibodies with the spike protein c. Affinity of the anti RBD/NTD antibodies as determined by BLI analysis

We next wanted to assess the assay’s sensitivity and specificity. To this end, recombinant spike and nucleocapsid proteins were both diluted (in PBS) and analyzed by the developed assay. Results, presented as signal-to-noise (S/N) values vs antigen concentration (Fig 2a and b, blue line), indicate that the assay recognizes the recombinant spike protein with no cross reactivity with the nucleocapsid. The assay enables detection of 1 ng/ml (S/N > 1.7, dashed line, Fig 2a) of the spike protein and its dynamic range spans three orders of magnitude.

**Fig. 2.**
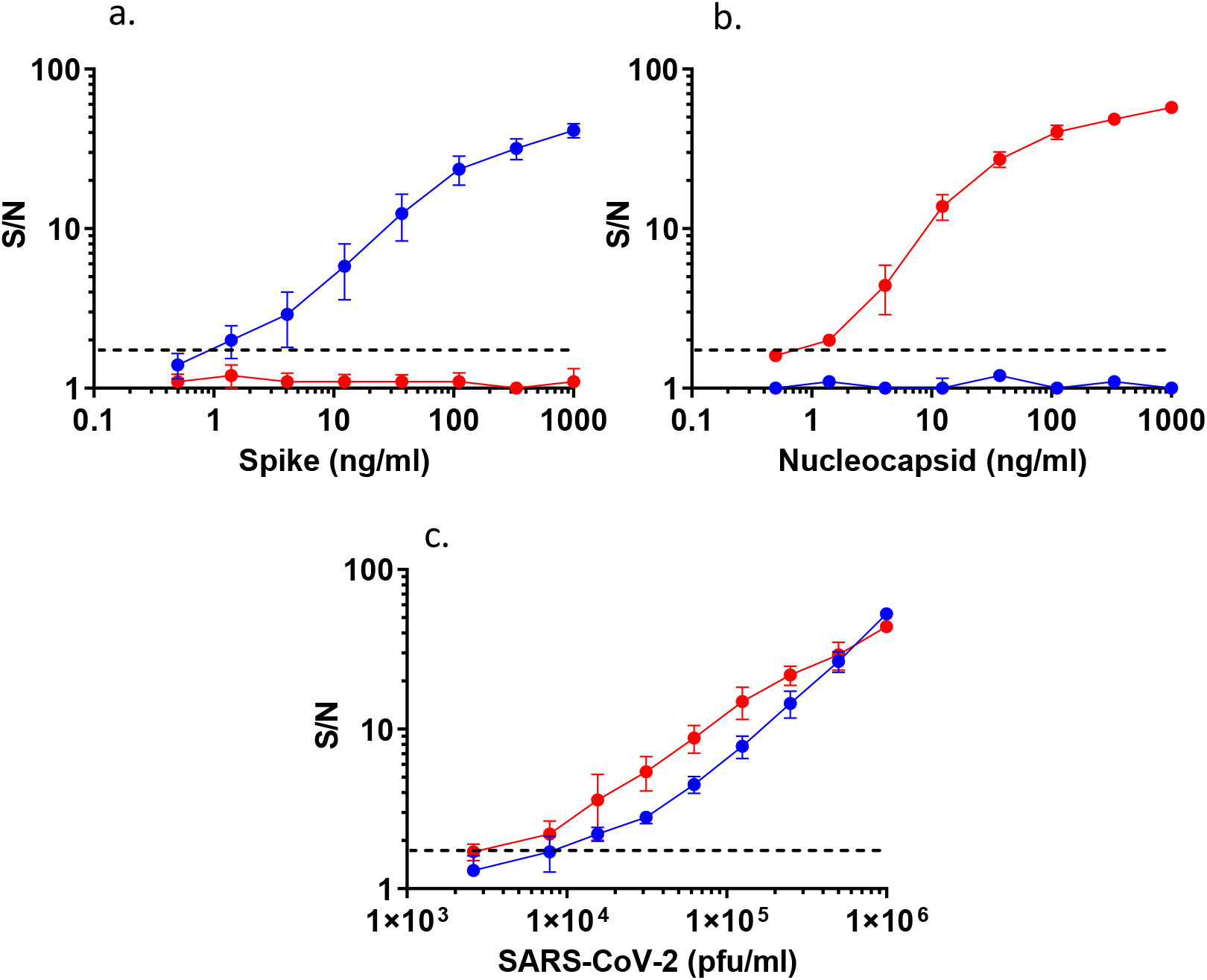
Dose response curves of TRF-based ELISA assays on recombinant antigens and SARS-CoV-2. Recombinant antigens: a. Nucleocapsid, b. spike or C. SARS-CoV-2 virus, were diluted in PBS (0.5-1000 ng/ml or 2×10^3^-1×10^6^ pfu/ml) and analyzed with the NC-assay (red) or S-assay (blue). Signal-to-noise (S/N) ratios were calculated as described in materials and methods. The dashed line represents the assay’s LOD

### Development of a TRF ELISA test for detection of SARS-CoV-2 nucleocapsid protein

In order to compare the sensitivity and specificity of SARS-CoV-2 detection via its spike to that based on the nucleocapsid, an additional in-house assay was developed. The nucleocapsid assay (NC-assay) is based on both polyclonal and monoclonal antibodies. The polyclonal antibodies, generated from a hyperimmune rabbit immunized with SARS-CoV-2 nucleocapsid, are implemented as capture antibodies. A monoclonal antibody (KD=0.7nM, kon=5.6×10^5^ 1/Ms; koff=3.8×10^−4^ 1/s), isolated from the same COVID-19 patient’s sera phage-display library, is applied as the reporter antibody. The NC-assay’s specificity and sensitivity were analyzed as described for the S-assay. Results (Fig 2a and b, red line) indicate that this assay specifically recognizes its intended target (nucleocapsid) with no detectable activity on the reciprocal antigen (spike). Similar to the S-assay, this assay also demonstrates a wide dynamic range (three orders of magnitude) and enables detection of 1 ng/ml of the recombinant nucleocapsid.

### Detection of cultured SARS-CoV-2

In the previous sections we described the development of two in-house assays enabling specific detection of their recombinant intended targets. We next applied both assays for SARS-CoV-2 detection using serial dilutions of propagated viral culture supernatants (Fig 2c). The assays facilitate detection of 1×10^4^ pfu/ml and 8×10^3^ pfu/ml SARS-CoV-2, for the S-assay and NC-assay, respectively. These results are equivalent to nasopharyngeal swab samples with cycle threshold (Ct) values of 25 and lower, according to a qRT-PCR test applied routinely for COVID-19 detection (24), indicating that both have the potential to be used for COVID-19 diagnosis.

### Detection of SARS-CoV-2 from nasopharyngeal swab specimens

Finally, we wanted to verify that the tests can be used for SARS-CoV-2 detection in nasopharyngeal swab specimens. To this end we tested a total of 146 qRT-PCR positive and 138 qRT-PCR negative swab samples. qRT-PCR analyzed swab samples were stored at 4°C (for up to a month), processed and tested in concert by both tests (not all samples were analyzed by both tests due to insufficient sample volume). The criteria for performance assessment were sensitivity, specificity and accuracy. In this respect, qRT-PCR results were used to determine “true positive” (meaning COVID-19 patient) or “true negative” (meaning healthy or non-COVID-19 patient) status for each sample. Evaluation was based on crude S/N ratios that were determined for each assay. A plot of S/N values vs Ct for both assays is presented (Fig 3). In this plot, Ct values above 41 represent negative qRT-PCR samples. S/N values were than translated to “positive” and “negative” results based on a calculated cut-off value (as detailed in materials and methods), resulting in contingency tables that were analyzed with GraphPad Prism 6 (Table 1). The analysis is presented separately for all the samples, including the full spectra of Ct values (“all Ct”, Table 1 left panel) and for samples with high viral loads (“Ct<25”, Table 1 right panel). Results (Table 1 and Fig 3) indicate that both assays are suitable for COVID-19 diagnosis from nasopharyngeal swabs. As indicated (Table 1), the S-assay demonstrates higher specificity (99.1%) compared to that of the NC-assay (87.0%). These results are in accordance with CDC’s lab alert, cautioning against false positive results when implementing antigen-based detection (which is mostly nucleocapsid-based). Alas, the higher specificity of the S-assay is coupled with lower sensitivity: 49.3% vs 63.4% for the S-assay and the NC-assay respectively (Table 1, left panel). The sensitivity of both assays improves when analyzing high viral load (Ct<25) samples (Table 1, right panel) to 65.7% and 84.5% for the S-assay and NC-assay respectively. The high sensitivity/low specificity vs low sensitivity/high specificity of both tests is clearly demonstrated by the shift in the positive results “cloud” of the NC-assay towards detection at higher Ct values (sector: 25<Ct<40, S/N>1.7, Fig 3a and b), while demonstrating a higher rate of false positive results for negative samples (sector Ct > 40, S/N>1.7, Fig 3a and b).

**Table 1:**
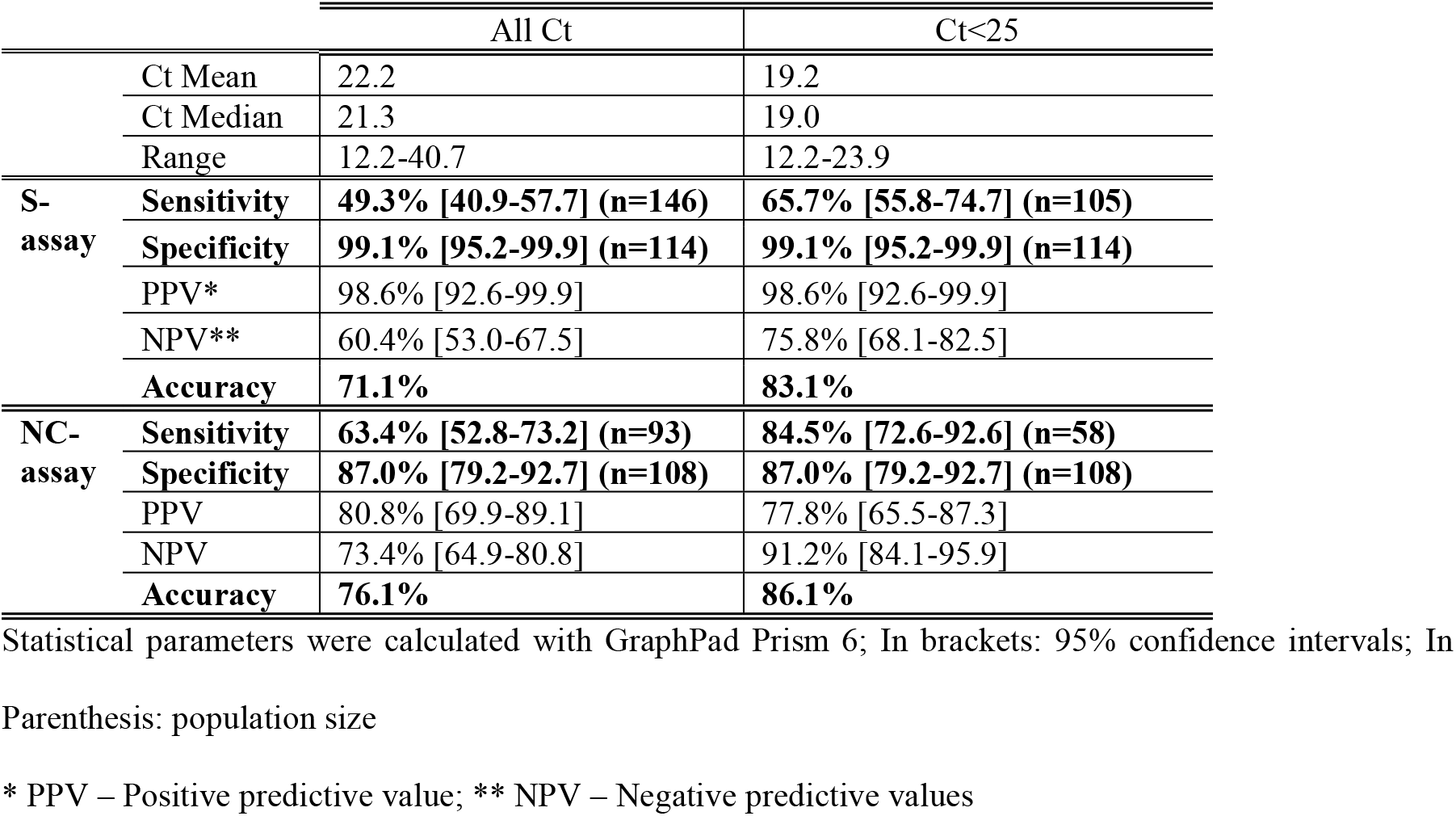
Clinical performance of TRF ELISA tests.

**Fig. 3.**
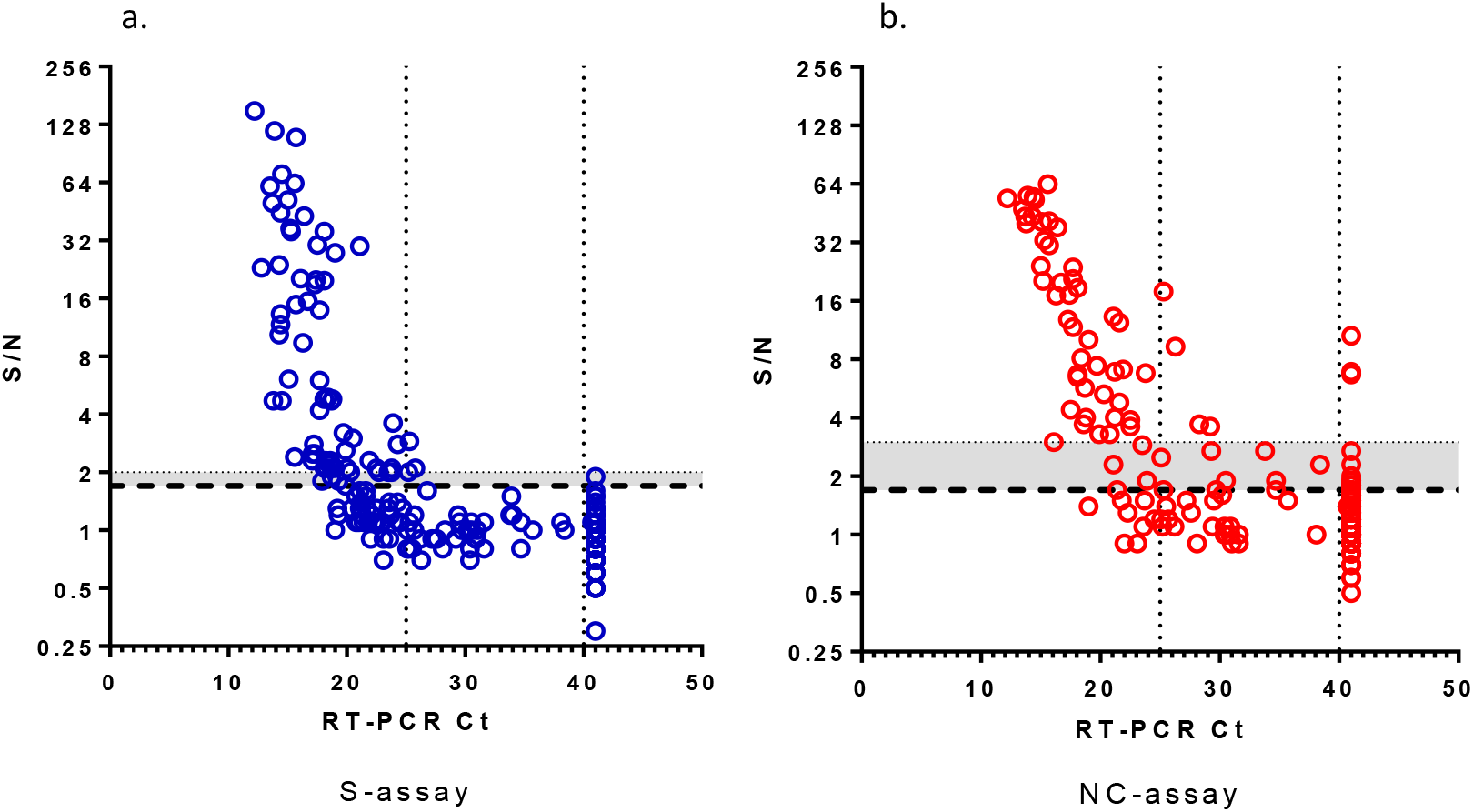
S/N ratios vs Ct values. a. S-assay b. NC-assay. The assay’s LOD is indicated by the dashed line. Ct value > 41 represent negative qRT-PCR samples. The gray area, enclosed by the dotted line, represents the difference in sensitivity required for enhanced specificity

We wanted to determine whether the difference in sensitivity between the tests is statistically significant (Fig 4a). A Kruskal-wallis multiple comparison test indicated that while the difference between Ct values of detected (Positive - P) vs undetected (False negative - FN) within each test is statistically significant (P values<0.001), the difference between tests is not statistically significant. This observation suggests that based on the relatively small number of samples analyzed in this study, the sensitivity of both tests is similar and a larger sample size is needed to determine a clear distinction. An attempt to improve the NC-assay specificity by elevating the cutoff value (Fig 3b, gray area) further lowers the difference between the assays sensitivity as this change is accompanied by a decline in assay’s sensitivity.

**Fig. 4.**
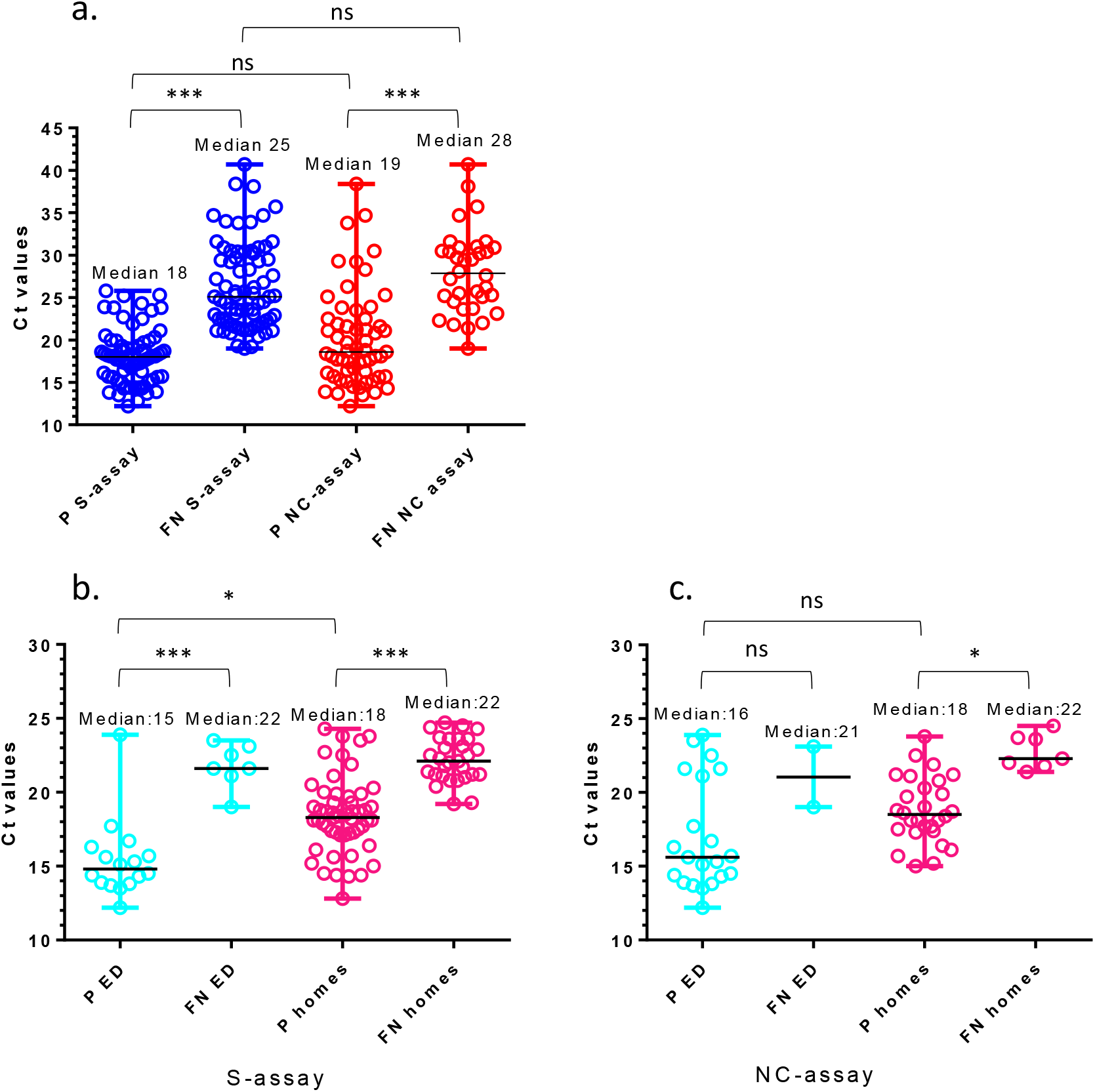
Comparison of viral loads (Ct values<25) among positive (P) and false negative (FN) results. a. For the S-(blue) vs NC-(red) assays (for all qRT-PCR positive samples). For qRT-PCR positive samples from emergency departments (ED) (cyan) and nursing homes (homes) (pink) utilizing b. S-assay; C. NC-assay. Statistical analysis was performed using GraphPad Prism 6, applying one-way ANOVA test followed by Kruskal-wallis multiple comparison tests. ns, not significant; * p <0.05; *** p <0.001

One of the main research questions examined in this study was whether assay performance depends on the scanned population i.e. whether there is a difference in between SARS-CoV-2 recognition in swab specimens originating from symptomatic vs asymptomatic people. This question is valid for both qPCR-positive and negative samples. To test a possible correlation, we re-evaluated the results in accordance with sample origin. Samples were collected either from routine screening of nursing homes (personnel and residents) or from the emergency department (ED) of a local hospital. While both positive and negative ED samples were mostly of symptomatic individuals (the samples were accompanied with information regarding the presence of symptoms and the time since symptom onset), the nursing homes samples had no additional information besides their qRT-PCR results. The assumption, that nursing homes samples include positive samples from asymptomatic or recuperating patients (several days after symptom onset and no longer infective), is reasonable. It is also logical to assume that most qRT-PCR negative samples originating from routine screening were probably collected from healthy individuals.

A total of 100 qRT-PCR positive and 115 qRT-PCR negative nursing homes samples and 46 qRT-PCR positive and 19 qRT-PCR negative ED samples were analyzed. The ED qRT-PCR positive population presented a Ct mean of 24.1, median of 24.5 and a range of 12.2 - 40.7. The nursing home samples presented a lower Ct mean of 21.3, median of 20.9 and a range of 12.8 – 34. A comparison of Ct values amongst positive (P) and false negative (FN) results from the emergency department (ED) and nursing homes (homes) was performed for both the S- and NC-assays (Fig 4b and c). Analysis was performed for high viral loads (Ct values<25) which are believed to indicate viable contagious virus (25, 26). A statistically significant difference (p<0.001) was observed between Ct values of PCR-positive samples detected (P) vs un-detected (FN) using the S-assay, regardless of the tested population (ED vs homes) (Fig 4b) (Kruskal-Wallis test performed using GraphPad Prism 6). No (ns) or low (p<0.05) difference were obtained for the same populations using the NC-assay (Fig 4c). However, as opposed to the NC-test, a statistically significant difference (p<0.05) was also observed for the S-assay, between the mean of the Ct values of detected (P) samples collected from the emergency department (P ED) to those collected from nursing homes (P homes) (Fig 4b). This difference might result from an actual variance between the Ct values presented by symptomatic vs a-symptomatic COVID-19 patients. This observation in not supported by the literature (25, 27) and cannot be deduced from such a small sample size (n=23). Another explanation for the difference might arise from the test itself. According to several publications, the viral loads in the upper respiratory tract peak around symptom onset and persists for 10 days in mild-to-moderate disease, with qRT-PCR Ct values strongly correlating with cultivable virus (25, 27). Since no information regarding days from symptom onset accompanied the nursing home samples, the presence of residual RNA (indicated by the qRT-PCR test), does not necessarily point to the presence of viable virus which in turn is accurately not detected by the assay (26). One can surmise that despite the statistical difference, assay performance is similar for both the ER and nursing homes samples and depends solely on the assay’s LOD.

## Discussion

Several commercial point-of-care (POC) devices were developed in an attempt to minimize the time gap between sampling and result delivery. These tests (28) are mainly based on immuno-detection of the encapsulated SARS-CoV-2 nucleocapsid protein. There are several publications presenting antigen detection tests based on the virion’s externally presented spike trimers (9-17), however, only one of the commercial tests approved by the FDA is based on spike detection (Sampinute COVID-19, Celltrion USA, Inc). Due to a certain rate of false positive results encountered when implementing some of the nucleocapsid-based commercial kits (as indicated by warnings issued by both the CDC and FDA), we thought it prudent to evaluate spike vs nucleocapsid SARS-CoV-2 detection. In this work we present a side-by-side comparison of two in-house ELISA-based assays for detection of SARS-CoV-2 from a large cohort of nasopharyngeal swabs targeting its spike or nucleocapsid antigens.

The assays presented in this study are ELISA-based and therefore may be easily applied in clinical laboratories. Moreover, the antibodies can be implemented in commercial point-of-care (POC) technologies, some of which demonstrate very high sensitivities (11, 14, 16). POC tests could be beneficial for two distinct possible scenarios (Fig 5). The first (Fig 5a) would be as a front-line screening methodology of symptomatic patients (for example at EDs). In this setup a very specific test would be needed, as the implication of a positive antigen result would be the hospitalization/quarantine of the subject in a corona ward. The sensitivity of this assay is not as crucial, since a negative test result of a symptomatic individual would require a follow up PCR test (with appropriate preventive measures taken in the meanwhile). The second scenario (Fig 5b) would be for routine screening of healthy/asymptomatic populations (for example in schools). In this case, the test ought to be as sensitive as possible even at the price of a lower specificity, as only positive individuals would be PCR tested. A positive result in this case probably indicates a high viral load, marking this person as one who is likely to spread the disease (29, 30). This individual would then be quarantined at home until the arrival of his PCR results, thus lowering the risk of disease spread. While a negative result in this setup does not necessarily indicate a healthy individual, it might hint at a lower viral load, which is possibly less likely to cause disease spread. The viral load is liable to either increase with disease progression, enabling detection during the next routine scan or decrease further, as part of the natural resolution of infection.

**Fig. 5.**
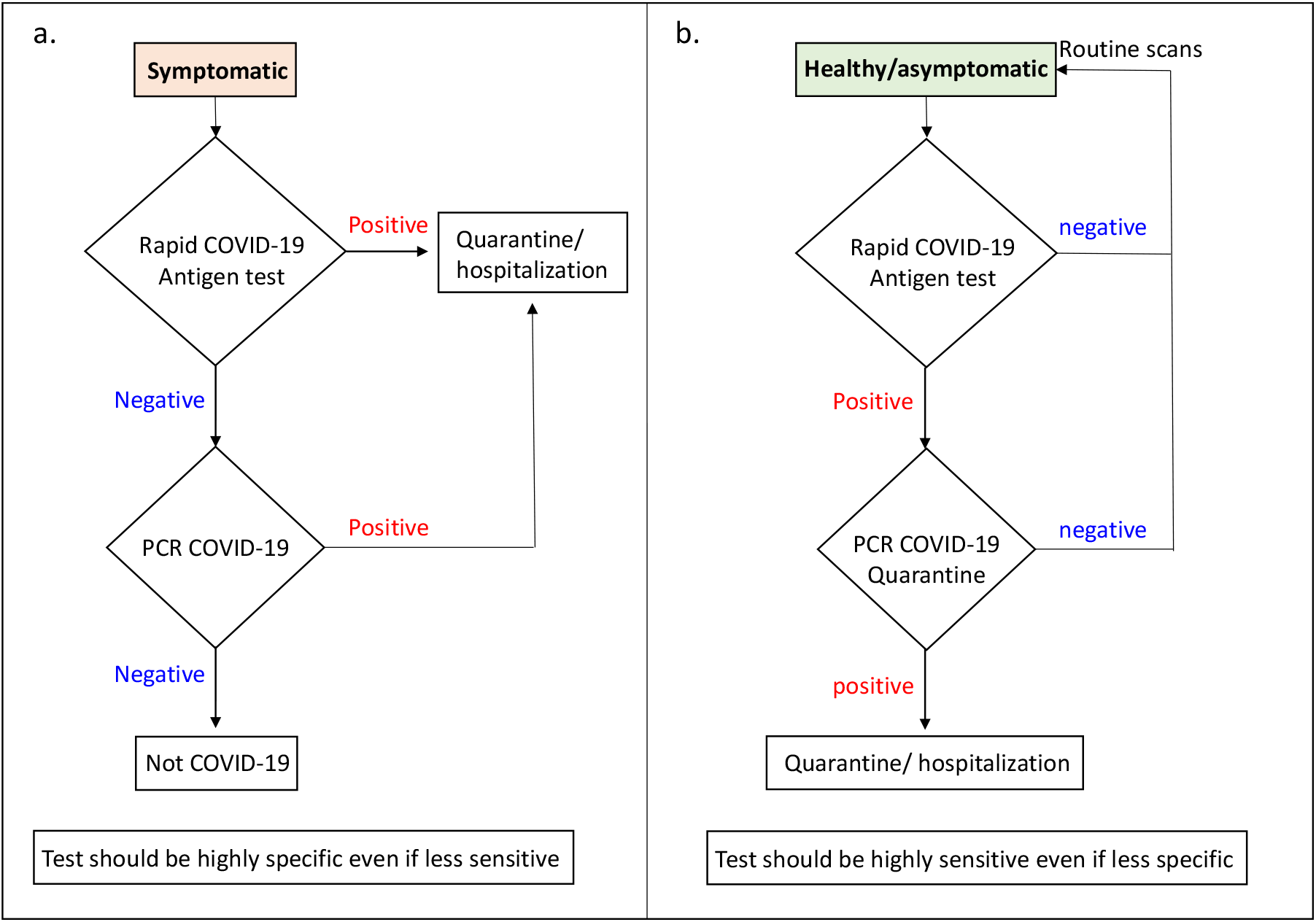
Logic flow chart for possible applications of immune-detection tests a. For screening of symptomatic patients. b. For routine screening of health/asymptomatic individuals

In the context of the suggested scenarios presented in Fig 5, it is important to note that in our hands the sensitivity of each test seemed to correlate directly with the viral load regardless of whether the sample originated from a symptomatic or an asymptomatic individual in accordance with the literature (25, 27). However, for both applications, antigen tests are suitable up to 5-10 days after disease onset while viral loads are high and the test actually indicates the presence of the virus and not residual RNA.

The antigen of choice in most commercial assays i.e. the nucleocapsid, indeed demonstrated higher sensitivity than that observed with the spike-based assay (Fig 3 and table 1). This higher sensitivity might have been predicted in advance, given that according to the literature, only around 100 spike trimers are present on each virion, resulting in an estimated total of 300 monomers, while around 1000 copies of the nucleocapsid are expressed in each virion (31). Non-specific reactions however, are harder to predict given that sequence homology is not the only indicator for possible interactions. In this regard, serological studies indicate that cross-reaction of pre-existing antibodies in SARS-CoV-2 negative individuals with the spike protein of endemic and seasonal corona viruses is low and is predominantly directed against S2 (32, 33). This might indicate that our S-assay, comprised of a combination of anti RBD and NTD antibodies, will exhibit high specificity. The fact that the spike assay is comprised of four different, high affinity, monoclonal antibodies can enhance specificity while still maintaining the ability to recognize emerging mutants (34). In another serological study (35), the authors examined cross-reactions of SARS-CoV-2 nucleocapsid with other coronaviruses in an attempt to assess its utility for seroprevalence studies. The authors found that anti-SARS-CoV-2-nucleocapsid IgG also recognized N229E nucleocapsid and IgG directed against HKU1 nucleocapsid recognized SARS-CoV-2 nucleocapsid. Cross reactivity was stronger with alpha-rather than beta-coronaviruses despite having less sequence identity, reiterating the importance of conformational recognition. Other coronaviruses notwithstanding, unpredicted interactions with bacteria or viruses present in healthy or symptomatic individuals might cause non-specific interactions (as is indeed the case for some of the commercial nucleocapsid-based antigen tests). It seems that only sampling of a large population of healthy or “corona-like” symptomatic individuals will provide quality data regarding true assay specificity (in addition to cross reactivity studies with known viruses, bacteria and fungus, as was performed by some manufacturers, for example CORIS BioCencept with COVID-19 Ag Respi-Strip). Our observations indicate that the spike-based test was significantly more specific than the nucleocapsid based test (Table 1).

The evaluated parameters of sensitivity and specificity depend solely on the cut-off values for positive vs negative results for each test. As indicated in Fig 3, the gray zone, delimited by the dashed and dotted lines, represents the decline in sensitivity resulting from elevating the cut-off value in order to improve assay specificity. We applied receiver operating characteristic (ROC) analysis and multinomial logistic regression in an attempt to define an optimal LOD for both tests. While the S-assay displays 100% specificity when applying a S/N ratio>2, for the NC-assay, in order to achieve 98% specificity, assay cut-off should be above S/N>7 (resulting in an increase in the assay’s LOD to 4×10^5^ pfu/ml). Application of different antibodies might improve assay’s specificity.

Another important issue when considering which antigen in more suitable for SARS-CoV-2 detection is the genetic variability and mutation rate of both antigens. During the evolution and global spread of the pandemic, this topic was addressed by several labs (36-38), describing several viral mutants that are now circulating all over the world (34). It is logical to assume that escape mutants are more likely to manifest in the spike protein (especially for mutants arising due to excessive use of plasma from recuperating individuals for treatment of COVID-19 patients), however researches discovered mutations in both genes (36-38). The nucleocapsid, as well as open reading frames 3a and 8, are implicated as having roles in virulence, transmission and pathogenicity (39) and are therefore as likely to undergo mutations as the spike protein despite not being presented on the virus surface. We therefore propose that for SARS-CoV-2 detection, both antigens will be used simultaneously or consecutively as was also suggested by others (40). The combination of both antigens on the same device might strengthen the reliability of the observed result.

## Data Availability

Data is available

## Declarations

## Ethics approval

The study was conducted according to the guidelines of the Declaration of Helsinki, and approved by the Institutional Review Board of Sheba Medical Center (protocol code 7769-20-SMC, Oct 19th, 2020).

## Consent for publication

All authors read and approved of the final manuscript

